# Occupationally exposed and general population antibody profiles to influenza A viruses circulating in swine as an indication of zoonotic risk

**DOI:** 10.64898/2026.01.08.26343691

**Authors:** Celeste A. Snyder, Garrett M. Janzen, Giovana Ciacci Zanella, Daniel C. A. Moraes, Gustavo S. Silva, Jefferson J. S. Santos, Elizabeth M. Drapeau, Scott E. Hensley, Tavis K. Anderson, Phillip C. Gauger, Amy L. Baker

## Abstract

Individuals with occupational exposure to swine may have disproportionate risk for zoonosis with swine influenza A virus (IAV). To evaluate human antibody responses, sera or plasma from swine veterinarians, swine farm employees, and the general population were tested by hemagglutination inhibition (HI) assays against representative swine and human seasonal influenza vaccine strains. HI data were analyzed by antigenic cartography to assess strain relationships, and reproduction number modeling to evaluate pandemic potential using age-stratified immunity profiles. Occupationally exposed groups had lower human seasonal vaccine uptake (45.5% vs 70%) and significantly lower odds of seropositivity to several H1 and H3 from swine compared to general population cohorts. One swine strain exhibited significant antigenic drift (3.62 AU) from its nearest vaccine strain. Multiple strains required lower R₀ thresholds for pandemic spread (1.09-1.35) than recorded pandemic strains (1.46-1.80). This demonstrates that population immunity gaps heighten zoonotic risk to circulating swine H1 and H3 strains.

## Introduction

Influenza A viruses (IAV), from the family Orthomyxoviridae, cause respiratory disease in humans and swine. Four influenza pandemics occurred from 1918 to present, with varying gene segments attributed to animal-origin IAV. The 1918 H1N1 virus was thought to arise from adaptations of avian influenza viruses (*1*). The 1957 H2N2 and 1968 H3N2 pandemics originated reassorted viruses containing gene segments derived from human seasonal and avian-origin IAV (*2*). The 2009 pandemic was a reassortment of viruses containing gene segments derived from avian, swine, and human IAV (*3*), and occurred when an H1N1 virus transmitted from pigs to humans before spreading among humans globally (*4, 5*). Viruses circulating in swine pose a threat to public health; IAV in swine resulted in 578 confirmed zoonotic cases in the United States (U.S.) since 2010 (*6*). Consequently, determining strains with zoonotic potential through measurements of genomic diversity, antigenic drift relative to human seasonal or candidate vaccine viruses, and human population immunity derived from vaccination or natural IAV infection is critical.

Commercial swine farms in the U.S. have endemic circulation of H1N1, H1N2, and H3N2 IAV. Each subtype has significant genomic diversity resulting from historic human-to-swine introduction of H1, H3, N1, and N2 gene segments that established unique lineages, along with H1N1pdm09 viruses (*7*). Currently, three lineages of H1 viruses are frequently detected in pigs globally: classical swine (1A), pre-2009 human-seasonal spillover (1B), and Eurasian avian origin (1C) viruses (*8*). Human seasonal H3 viruses are regularly introduced into swine populations since the 1968 H3N2 human pandemic, with lineages grouped by the decade of the human-to-swine introduction (*9*). As swine are susceptible to both avian and human IAVs, in addition to endemic swine-adapted strains, reassortment may result in novel genotypes with increased zoonotic potential (*10, 11*).

The evolution of IAV in swine results in antigenically drifted viruses, to which the human population may have little to no prior immunity (*12, 13*). Immune profiles to IAV derived from vaccination or exposure to IAV are not homogenous. Numerous demographic factors, including age, sex, ethnicity, income, and health insurance status, significantly influence influenza vaccination rates among adults in the U.S., subsequently impacting their potential protection against the virus (*14*). Veterinarians were shown to have an increased risk of infection with many zoonotic pathogens(*15*). Additionally, farm employees working at the human-swine interface may have a heightened risk for zoonoses (*16–18*).

Understanding human population immunity to endemic IAV in swine, particularly among occupationally exposed individuals with regular swine contact, provides critical input for zoonotic risk assessments. This study quantified antibody response within different human populations to the most frequently detected strains of IAV in U.S. swine to evaluate if occupation or other demographic factors increased zoonotic risk. Representative swine antigens from the most frequently detected IAV HA clades in U.S. swine were tested against human sera derived from two occupationally exposed cohorts collected in the U.S.A., swine veterinarians and swine farm employees, and two general population cohorts from the U.S.A. and Hong Kong, China. Results revealed that humans lack neutralizing antibodies to many circulating clades of IAV in swine, heightening zoonotic risk. Notably, occupational groups were less likely to have positive HI titers compared to general population cohorts. Profiling human antibody titers against circulating IAV in swine highlights immunologic gaps across populations and gives early identification of high-risk strains prior to zoonoses.

## Materials and Methods

### Human Cohorts

Human sera or plasma (n=186) were collected to assess antibody cross-reactivity to IAV in swine. Occupational cohort one was comprised of 51 veterinarians attending a swine veterinary conference in the Midwestern U.S., self-identified as primarily swine-focused, and contributed sera in fall of 2021. This cohort was labeled “Veterinarian.” Occupational cohort two was comprised of 47 employees from five swine farms in the U.S. Midwest region and contributed sera in 2022 as the farms were enrolled. This cohort was referred to as “Farm Employee.” Veterinarian and Farm cohorts were asked to respond to a questionnaire including primary state, age, sex, time since last contact with swine, and if they received an influenza vaccine in the last 12 months. Cohort three from the general population included 40 participants from a vaccination study at the University of Pennsylvania in Philadelphia, PA, USA, sampled in 2021. This cohort was labeled “Philadelphia.” Plasma was collected 28 days post-vaccination.

To compare to a separate geographic region without circulation of the U.S. swine viruses, cohort four included sera collected in 2022 from 48 individuals from an observational population immunity study at the University of Hong Kong School of Public Health, in Hong Kong, China, and was labeled as “Hong Kong.” Data on age or decade of birth, sex, and vaccination status were provided for both Philadelphia and Hong Kong general population cohorts. Written informed consent was obtained from all participants. The study protocols were approved by the institutional review boards of the University of Hong Kong, University of Pennsylvania, and Iowa State University.

### Representative IAV in swine and reference human seasonal virus selection

IAV strains were selected to represent the genetic diversity of viruses circulating in the months prior to human sample collection. All available swine HA gene sequences were downloaded from the octofludb between July 2021 and June 2022 (*19*). These data represented 686 H3 HA and 1,294 H1 HA genes, derived from 25 U.S. states wherein 98.2% of the U.S. swine population resides (*20*). The data were aligned using default settings in mafft v.7.526 (*21*). Maximum likelihood HA gene trees for the H1 and H3 were inferred using IQ-Tree v.2.2.2 (*22*) following automatic model selection with statistical support assessed using single branch tests and the ultrafast bootstrap algorithm (*23*). For each of the identified genetic clades of IAV, a single selection was made using PARNAS v0.1.6 (*24*), and viruses were requested from the USDA IAV in swine surveillance system repository at the National Veterinary Services Laboratories for use in the antibody assays (*19*). The selection included 7 H1 viruses from two lineages, including four from the H1 1A classical swine lineage (1A.1.1.3, 1A.3.3.2 (H1N1pdm09), 1A.3.3.3-c1, and 1A.3.3.3-c3) and three from the H1 1B human-seasonal lineage (1B.2.1, 1B.2.2.1, and 1B.2.2.2) (Table 1). Three H3 viruses were selected from three lineages detected in U.S. swine: 1990.4.a, 2010.1, and 2010.2 (Table 1). The inferred phylogenetic trees were down-sampled for visualization using smot v.0.17.4 (*25*). The smot monophyletic sampling algorithm was applied, maintaining a randomly selected proportion of the HA genes within each clade (H1, 16%; H3, 32%). The code and data used in these analyses is provided at https://github.com/flu-crew/datasets. The virus antigen panel included contemporary H1 and H3 human seasonal influenza vaccine strains (HuVac) derived from the WHO-recommended 2020–2021 Northern Hemisphere influenza vaccine strains and a pre-2009 H1 vaccine strain that shared a common ancestor with the tested swine H1 1B strains.

**Table 1.** Representative IAV antigens from swine and human seasonal influenza vaccines (HuVac) were included in hemagglutination inhibition (HI) assays.

| Strain | Subtype | HA clade | NA clade | Abbreviation |
| --- | --- | --- | --- | --- |
| rg-A/Hawaii/70/2019 | H1N1 | 6B.1A.5a.1 | B.2 | HI19.HuVac |
| A/swine/Illinois/A02524514/2020 | H1N2 | 1A.1.1.3 | N2-1998B | IL20.1A.1.1.3 |
| A/swine/Iowa/A02524480/2020 | H1N1 | 1A.3.3.2 | N1-pdm | IA20.1A.3.3.2 |
| A/swine/Indiana/A02636365/2022 | H1N1 | 1A.3.3.3-c1 | N1-classical | IN22.1A.3.3.3-c1 |
| A/swine/Minnesota/A02635908/202 | H1N1 | 1A.3.3.3-c3 | N1-classical | MN21.1A.3.3.3-c3 |
| A/Beijing/262/1995 | H1N1 | NA | NA | BJ95.HuVac |
| A/swine/Iowa/A02635863/2021 | H1N2 | 1B.2.1 | N2-1998B | IA21.1B.2.1 |
| A/swine/Iowa/A02478968/2020 | H1N2 | 1B.2.2.1 | N2-2002B | IA20.1B.2.2.1 |
| A/swine/Iowa/A02524534/2020 | H1N2 | 1B.2.2.2 | N2-LAIV-98 | IA20.1B.2.2.2 |
| A/Hong Kong/45/2019 | H3N2 | 3C.2a1b.1b | A.2.2 | HK19.HuVac |
| A/swine/Iowa/A02635890/2021 | H3N2 | 1990.4.a | N2-2002B | IA21.1990.4.a |
| A/swine/Kansas/A02245675/2020 | H3N2 | 2010.1 | N2-2002A | KS20.2010.1 |
| A/swine/Indiana/A02635878/2021 | H3N2 | 2010.2 | N2-2016 | IN21.2010.2 |

NA=not applicable.

### Serology

Sera and plasma were treated with receptor-destroying enzyme (RDE) (Sigma-Aldrich, MO, USA) and heat-treated at 37°C for 18-20 hours. RDE was then inactivated at 56°C for 30 min. Sera was absorbed using packed turkey red blood cells (RBC) at 0.5% for 1 hour to remove nonspecific agglutinins. Representative swine strains from lineages H1 1A, H1 1B, H3 1990.4, and H3 2010.x, and representatives of human seasonal influenza vaccines (H1B historic human seasonal vaccine strain) were expanded in Madin-Darby canine kidney cells (MDCK) and tested against human serum and plasma according to established hemagglutination inhibition (HI) methods (*26*). In the analysis, geometric mean titers (GMT) were calculated by the log2 transformation of reciprocal titers divided by 10. HI titers of <10 were given a pseudo count 10 to permit log2 transformation. A reciprocal HI titer of ≥40 or a log2 scale of 2 was considered positive and a correlate of protection.

### Antigenic Characterization

Antigenic maps were generated with HI assay data using the Racmacs tool (*27, 28*) in R version 4.4.1. Euclidean distances between pairs of antigens were used to determine the antigenic distances of selected IAV circulating in swine to human seasonal vaccine strains. Antigen-serum distances were calculated to determine the proportion of individuals with a significant antigenic distance from circulating swine strains. One antigenic unit (AU) equals a two-fold difference in HI titer. An antigenic distance of ≥3 AU (8-fold difference in HI titer) was considered significant and is used as a threshold to indicate a potential need to revise components within the human seasonal influenza vaccine (*29*).

### Odds Ratio Analysis

To evaluate how occupation, birth year, sex, and vaccination status influenced the odds of seropositivity (reciprocal HI titer ≥40), odds ratios (OR) were conducted using cohort type (occupational exposure and general population), born 1946-1977 and 1978-2003, sex, and vaccination status. Birth decade was not included due to small sample sizes within individual decades, though descriptive analysis of seropositivity by birth decade is presented in Appendix Figure 1. The birth year cut off 1977 represents the re-emergence of H1N1 following the H2N2 and H3N2 pandemics. OR analyses were calculated in R version 4.4.1 with the epitools package (*30*), using median-unbiased estimation and Fisher’s exact methods for confidence interval calculation. Confidence interval calculations were not calculated for contingency tables containing one or more values of zero. OR analyses were carried out for composite groups of occupational exposure (Veterinarian + Farm Employee) and general population (Philadelphia + Hong Kong) to increase sample size and improve precision.

**Figure 1.**
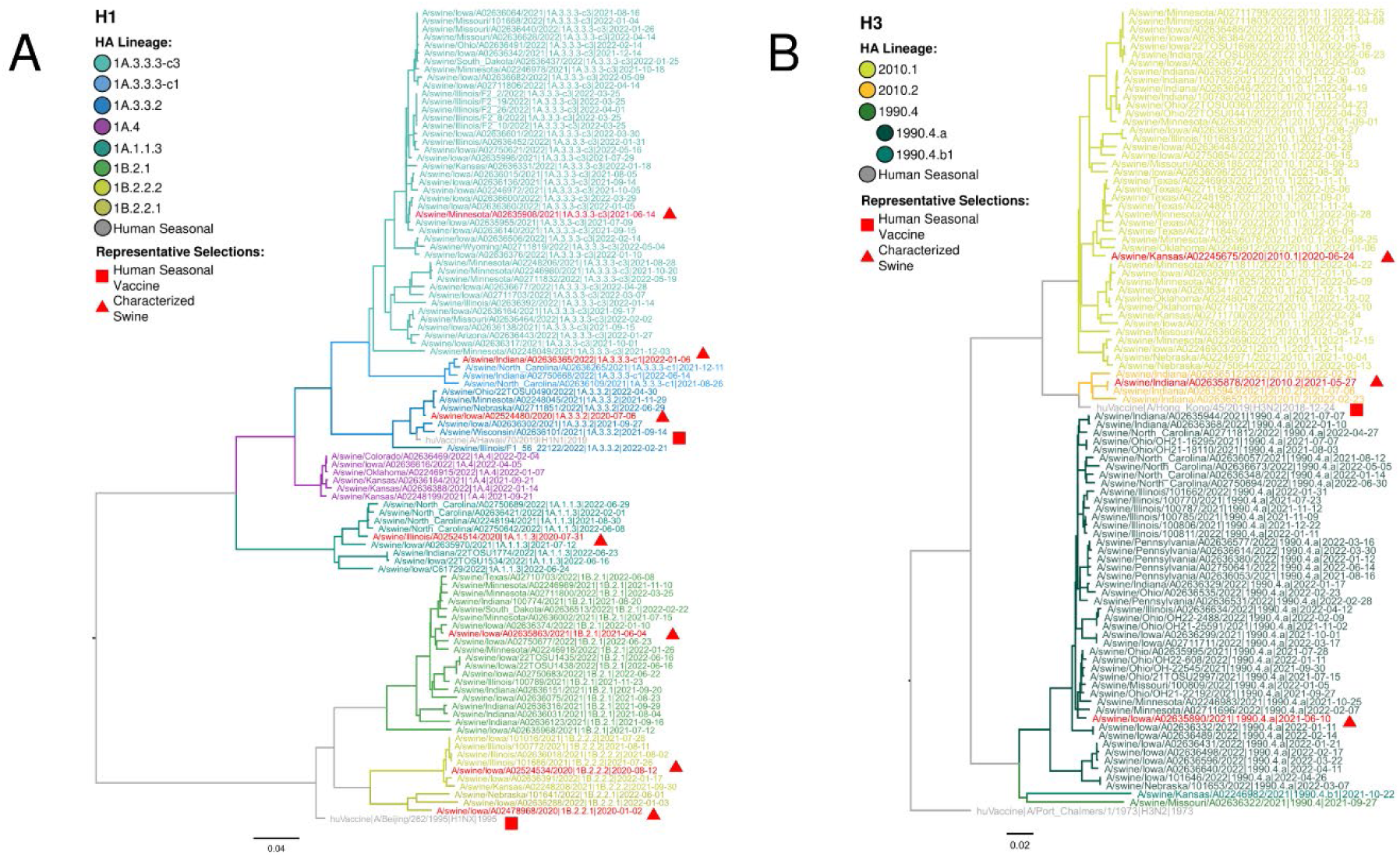
Representative maximum-likelihood phylogeny of (A) 645 swine and human H1 and (B) 341 swine and human H3 hemagglutinin (HA) genes. (A) The two major H1 HA lineages in U.S. swine resulted from independent introductions of H1N1 and H1N2 from humans to swine and are grouped by the 1A and 1B lineage designations. Each lineage is divided into and colored by statistically supported clades: 1A.1.1.3, 1A.3.3.2, 1A.3.3.3-c1, 1A.3.3.3-c3, 1A.4; and 1B.2.1, 1B.2.2.1, and 1B.2.2.2. Triangles and squares annotate selected reference and test antigens; human seasonal H1 HA genes are colored gray. (B) Three major H3 HA lineages in U.S. swine were derived from interspecies transmission in the 1990s (1990.4.x) and 2010s (2010.1 and 2010.2). The H3 tree was rooted on the human seasonal HA gene A/Port Chalmers/1/1973 (H3N2). Branch lengths were drawn to scale, and the scale bar indicates the number of nucleotide substitutions per site. The phylogeny with tip labels included is available at https://github.com/flu-crew/datasets.

### Modeling R₀ thresholds from serological data

Reproduction number (R₀) threshold estimates were calculated using age-stratified HI data from combined cohorts into 7 age groups (0-19, 20-29, 30-39, 40-49, 50-59, 60-69, 70+ years). This approach models the theoretical minimum viral transmissibility requirements for pandemic emergence based on existing population immunity gaps, rather than measuring actual viral basic R₀ values from transmission data. Reciprocal titer values ranged from <10 to 10,240, with values <10 assigned a value of 5. Parameters were derived from previously described studies (*31*). Values were estimated with 500 Monte Carlo bootstrap simulations, incorporating a previously described empirical contact matrix, aggregated to align with age stratification (*32*), and adaptive titer-protection relationships. Protection levels ranged linearly from 0% to 95% across titer levels, following prior pandemic risk assessment approaches (*33*). This approach estimated the population immunity proportion, relative R₀ reduction, GMT, and minimum R₀ threshold required for pandemic emergence. The R0 threshold reflects the minimum inherent viral transmissibility needed to overcome existing population immunity and achieve sustained human-to-human transmission.

## Results

### Human Cohorts Demographics

The Veterinarian cohort (n=51) included 38 (74.5%) males and 13 (25.5%) females and a median age of 43. In the self-reported questionnaire, 31 (60.8%) participants reported receiving the human seasonal influenza vaccine within the 12 months prior to sample collection, 19 did not, and 1 reported unknown. The cohort reported diverse residences across Iowa, Kentucky, Illinois, Indiana, Nebraska, Missouri, Minnesota, North Carolina, Pennsylvania, and Kansas. The Farm cohort (n=47) included five swine farms from the U.S. Midwest region in Iowa, Illinois, and South Dakota. Demographics included 29 (61.7%) males and 18 (38.3%) females with a median age of 37. Farm employees self-reported that 14 (29.8%) received the human seasonal influenza vaccine within the 12 months prior to sample collection, 31 did not, and 2 reported unknown. Individuals in the Farm and Veterinarian cohorts were considered to have occupational exposure with 72% (n = 98) reporting their last contact with swine within one day.

The Philadelphia cohort (n=40) included 17 (42.5%) males and 23 (57.55) females with a median age of 33. All (100%) individuals of this cohort received the 2021-2022 human seasonal influenza vaccine. The Hong Kong cohort (n=48) included 16 males (33.3%) and 32 females (66.7%) with a median age of 46 years old; 16 (33.3%) reported receiving vaccination, and 32 reported no vaccination the prior year.

### Genetic diversity of H1 and H3 IAV in swine

Between July 2021 and June 2022 during the human blood collection period, 8 H1 1A, 3 H1 1B, and 6 H3 clades from three lineages (1990.4, 2010.1, 2010.2) were detected from 25 U.S. states. The H1 subtype had significantly more detections over the period than the H3 subtype, with 645 sequences (65.4%) compared to 341 sequences (34.6%), respectively. The 1A.1.1.3 (10.6%), 1A.3.3.2 (7.1%), 1A.3.3.3-c3 (24.7%), and 1B.2.1 (15.7%) were the most frequently detected clades and collected in 25 U.S. states (Appendix Figure 2) (*19*). The H3 1990.4 lineage diversified to include four genetically distinct groups, but only the 1990.4.a (13.6%) represented more than 1% of detections. The H3 lineages, 2010.1 and 2010.2, represented 14.5% and 1.1% of the detections, respectively. We objectively selected 10 representative HA genes (Figure 1) that represented 97.8% of the detections in the USDA IAV in swine passive surveillance system between July 2021 and June 2022 (*19*). Strains containing the 10 selected HA genes used in the antibody assays, along with the human seasonal vaccine strains, are summarized in Table 1.

**Figure 2.**
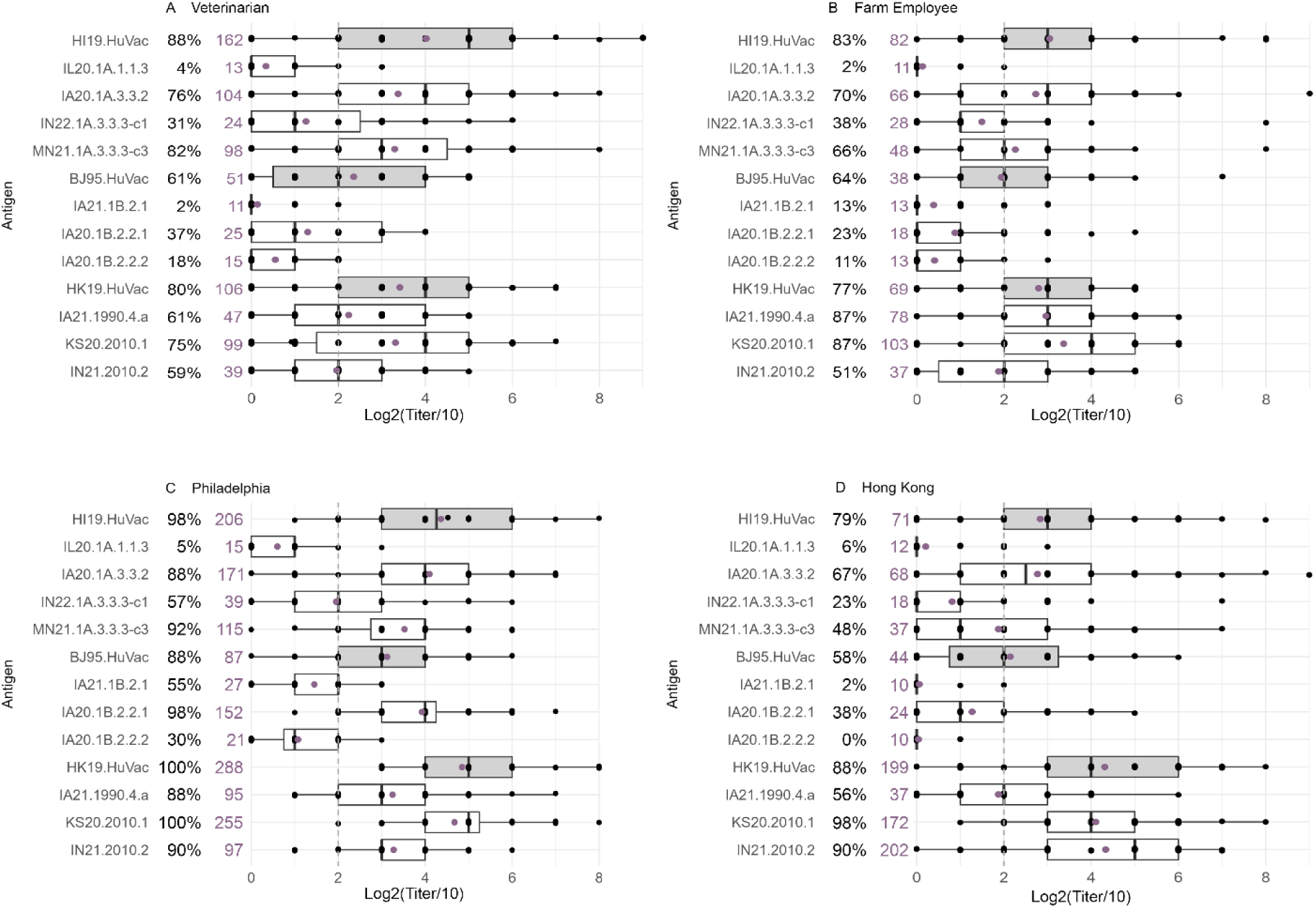
Box and whisker plots of log2 transformed hemagglutination inhibition (HI) titers among participants in four human cohorts against contemporary swine H1 and H3 influenza A virus strains. Box plots show median Log2 transformed HI titers on the x-axis, test strains on the y-axis, interquartile ranges, and outliers in A) Veterinarian (n=51), B) Farm Employee (n=47), C) Philadelphia (n=40), and D) Hong Kong (n=40). Purple dots represent the geometric mean log2 transformed HI titers/10. Gray box plots represent responses against human seasonal influenza vaccine strains, and white box plots represent responses against swine strains. Reverse-transformed HI geometric mean titers in purple and percentages of positive individuals in black are shown next to virus names on each panel. The gray dotted line indicates the HI titer threshold of 40 (log2 scale of 2).

### Antibody levels to IAV in swine by cohort

Titers of neutralizing HI antibodies to representative swine strains and human seasonal vaccine strains were generated for each human participant. These data demonstrated swine IAV strains with patterns of low HI responses in occupational and general population cohorts. Titers against IA20.1A.3.3.2 and MN21.1A.3.3.3-c-3 were higher compared to titers against IL20.1A.1.1.3, IN22.1A.3.3.3-c-1, IA21.1B.2.1, and IA20.1B.2.2.2 for all cohorts (Figure 2). Although swine H3N2 representatives elicited higher GMT and percentages of seropositive individuals overall, 10.2–28.5% of participants fell below the positive cutoff (Log2[titer/10] or 40 GMT).

In the Veterinarian cohort, 88% of individuals were HI positive with a GMT=162 for the human H1N1 vaccine component, HI19.HuVac, 61% of individuals were HI positive with GMT=51 for the pre-2009 historical H1N1 vaccine strain BJ95.HuVac, and 80% HI positive with GMT=105 for the human H3N2 vaccine component, HK19.HuVac (Figure 2A). Similarly, 83% of individuals in the Farm cohort were HI positive with a GMT=82 for HI19.HuVac, 64% of individuals were HI positive with GMT=38 for strain BJ95.HuVac, and 77% HI positive with GMT=68 for HK19.HuVac (Figure 2B).

The Philadelphia cohort displayed the highest overall antibody response with 98% HI positivity for H1N1 HI19.HuVac GMT=206, 88% for H1N1 BJ95.HuVac GMT=87, and 100% for H3N2 HK19.HuVac GMT=288 (Figure 2C). HI results for contemporary swine strains clades 1A.1.1.3, 1A.3.3.3c-1, 1B.2.1, and 1B.2.2.2 showed broader antibody cross-reactivity than in other cohorts, although some individuals in every cohort were seronegative (Figure 2C). In the Hong Kong cohort, 79% of individuals were HI positive with a GMT=71 for HI19.HuVac, 58% of individuals were HI positive with GMT=44 for BJ95.HuVac, and HK19.HuVac had 88% HI positive with GMT=199 (Figure 2D). This cohort followed similar trends to the other cohorts, except titers to IN21.2010.2, which displayed higher GMTs and percent positives similar to the human seasonal vaccine strain HK19.HuVac (Figure 2D).

### Impact of demographics and vaccination on seropositivity patterns

Demographic factors significantly impacted patterns of seropositivity across all IAV strains tested (Figure 3). Occupational exposure to swine was associated with significantly lower odds of seropositivity for H1 1B (OR = 0.23, 95% CI: 0.11-0.48, p < 0.001) and H3 lineages (OR = 0.17, 95% CI: 0.05-0.44, p < 0.001) compared to general population cohorts. Uptake of the human seasonal vaccine produced the strongest seropositivity association across all subtypes, H1 1A (OR = 5.30, p < 0.001), H1 1B (OR = 5.48, p < 0.001), and H3 (OR = 14.44, p < 0.001).

**Figure 3.**
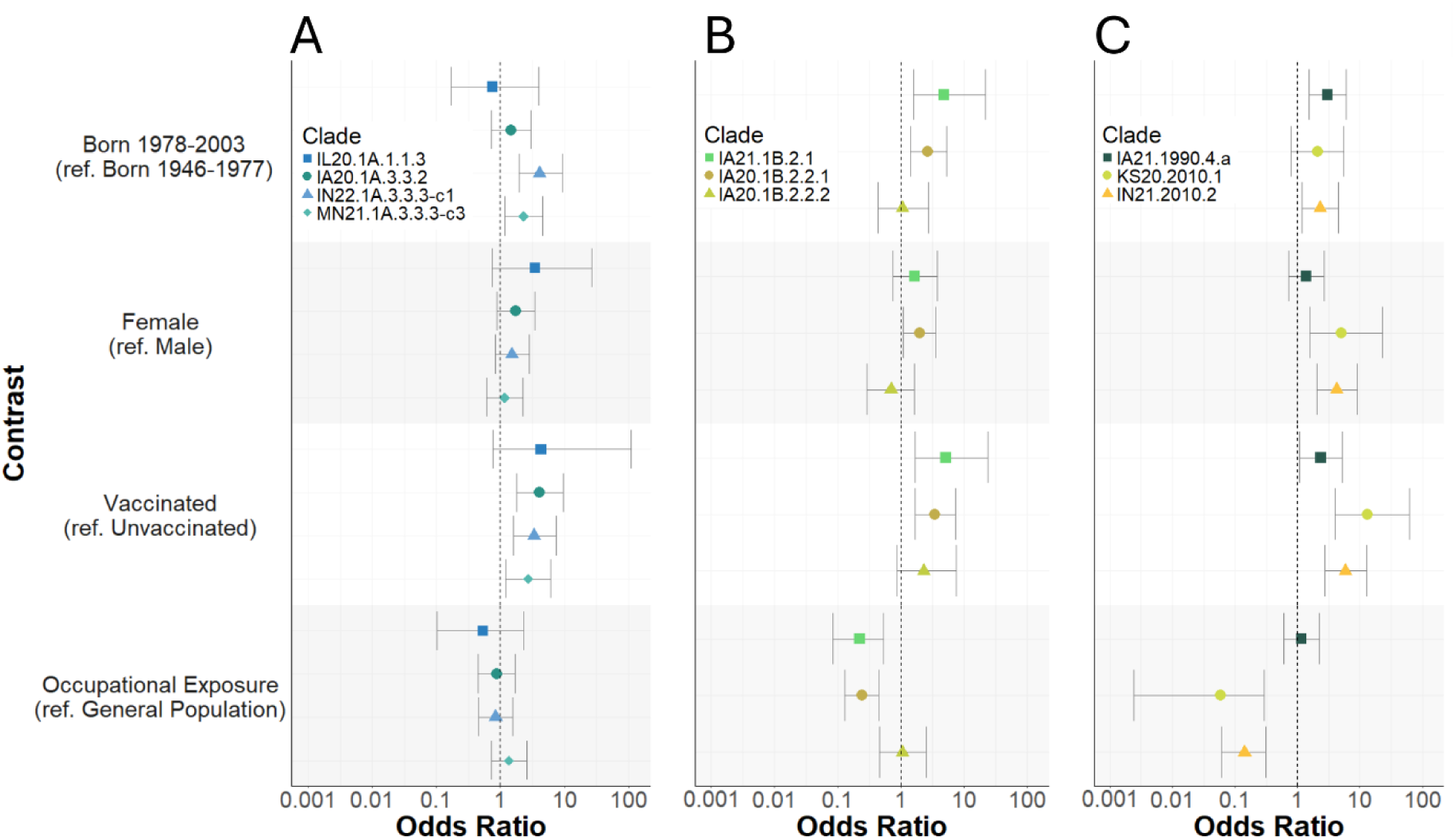
Odds ratios and 95% confidence intervals for seropositivity for IAV in swine clades (A) H1 1A, (B) H1 1B, and (C) H3 for demographic and vaccination status variables. References are noted in parentheses for each variable on the y-axis. Odd ratios displayed on logarithmic x-axis scale.

Additionally, individuals born from 1978-2003 showed higher seropositivity than those from 1946-1977, for H1 1A (OR=1.94, p=0.038), H1 1B (OR = 3.47, p = 0.002), and H3 lineages (OR = 2.35, p = 0.036) (Appendix Table 1).

At the strain level, tested H1 1B strains showed vulnerabilities in the occupational cohorts across the lineage (OR = 0.23), with IA21.1B.2.1 and IA20.1B.2.2.1 demonstrating strong negative associations (ORs 0.22-0.24, both p < 0.001) (Appendix Table 3). H3 lineages displayed the strongest overall negative association with occupational exposure (OR = 0.16), though this was driven primarily by KS20.2010.1 (OR = 0.06, p < 0.001) (Table S4).

### Antigenic distance of IAV in swine to the representative human seasonal vaccine strains and seropositive individuals

Antigenic maps were used to visualize HI data, independent of vaccination status or other demographic factors, relative to the nearest human seasonal vaccine strain and displayed for each phylogenetic lineage of the swine strains (Figure 4). In the H1 1A lineage, the swine representative of the clade originating from the human 2009 H1N1 pandemic (1A.3.3.2) was the most closely related antigen to the human vaccine strain HI19.HuVac (0.39AU). The 1A.1.1.3 strain displayed the highest antigenic distance from HI19.HuVac (3.62 AU), with 69% of individuals with significant antigenic distance >3AU (Figure 4A). However, the association between prior vaccination and responses to this strain could not be reliably assessed due to low seropositivity (Appendix Table 2). Clade 1A.3.3.3-c1 displayed 31% of individuals with a significant antigenic distance (2.54 AU) (Figure 4A). In the H1 1B lineage, the 1B.2.2.1 strain was the most closely related antigen to the historic human seasonal vaccine strain BJ95.HuVac (1.07 AU). Clade 1B.2.1 and 1B.2.2.2 strains had 22% and 19%, respectively, of individuals exhibiting significant antigenic distance from the swine strains (Figure 4B). H3 lineage antigens displayed relatively minimal antigenic distance compared to the human seasonal vaccine strain HK19.HuVac. However, 10% of individuals had significant antigenic distance to the 1990.4.a clade antigen (2.17 AU) (Figure 4C).

**Figure 4.**
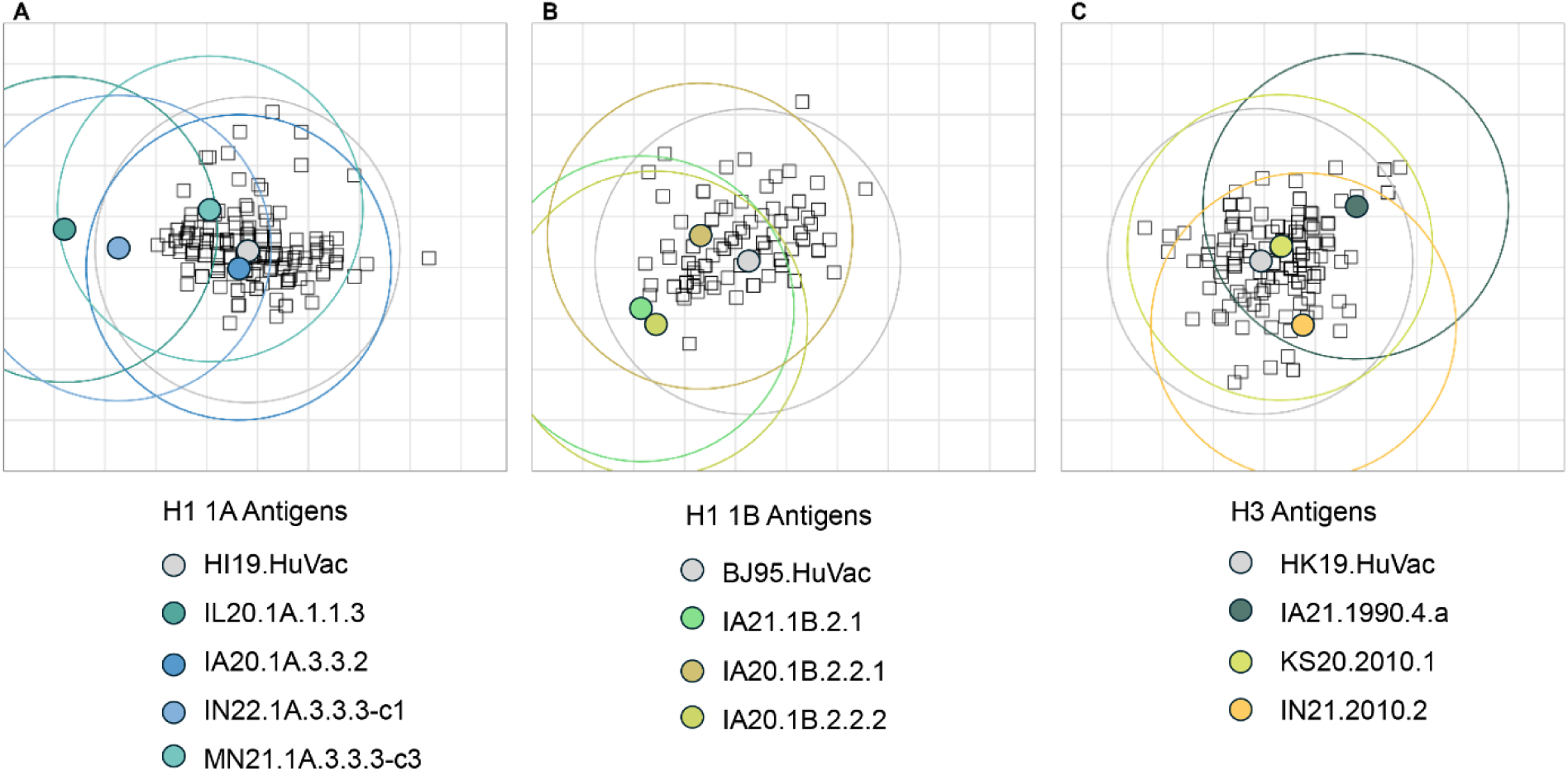
Antigenic map showing the relationships between human antibody immunity and selected strains of IAV in swine and human seasonal vaccines. Each sphere on the map represents an antigen, while squares represent individual participant serum samples from the combined Veterinarian, Farm, Philadelphia, and Hong Kong cohorts. (A) H1 1A classical swine lineage derived from the 1918 human influenza pandemic. (B) H1 1B human seasonal lineage derived from introductions of pre-2009 human seasonal H1 IAV. (C) H3 lineages derived from decadal introductions of human seasonal H3N2 viruses into swine. Rings around each strain display 3 antigenic units (AU) or 8-fold change in titer from the associated antigen and individuals outside the ring are predicted to have significant loss in cross-protection.

### Assessment of pandemic potential from population immunity

Since occupational and general population categorization was not a significant indicator of HI seropositivity, individuals were combined to give better distribution across the age classes. Reproduction number modeling revealed that pandemic vulnerability based on human population immunity ranged from 16.3% to 52.5% across tested antigens, with minimum R₀ thresholds required for pandemic emergence ranging from 1.09 to 2.36 among the swine strains (Figure 5). Four swine strains demonstrated low R₀ pandemic thresholds, indicating the potential to achieve pandemic spread by overcoming population immunity, with lower minimum inherent human transmissibility than the 2009 H1N1 pandemic virus (minimum R₀ < 1.46) (*34*). The H1 1A strains had low to moderate R₀ thresholds (range: 1.09-1.83). All H1 1B lineage strains showed low thresholds (range: 1.11-1.35), while H3 strains had relatively higher R₀ values (range: 1.93-2.36). The lowest R₀ thresholds were observed for IL20.1A.1.1.3 (minimum R₀ = 1.09, 95% CI: 1.07-1.28) and IA21.1B.2.1 (minimum R₀ = 1.11, 95% CI: 1.09-1.30), with only 16% of individuals demonstrating immunity (Appendix Figure 3), indicating these swine strains have the potential to emerge in the human population even with relatively low inherent transmissibility requirements.

**Figure 5.**
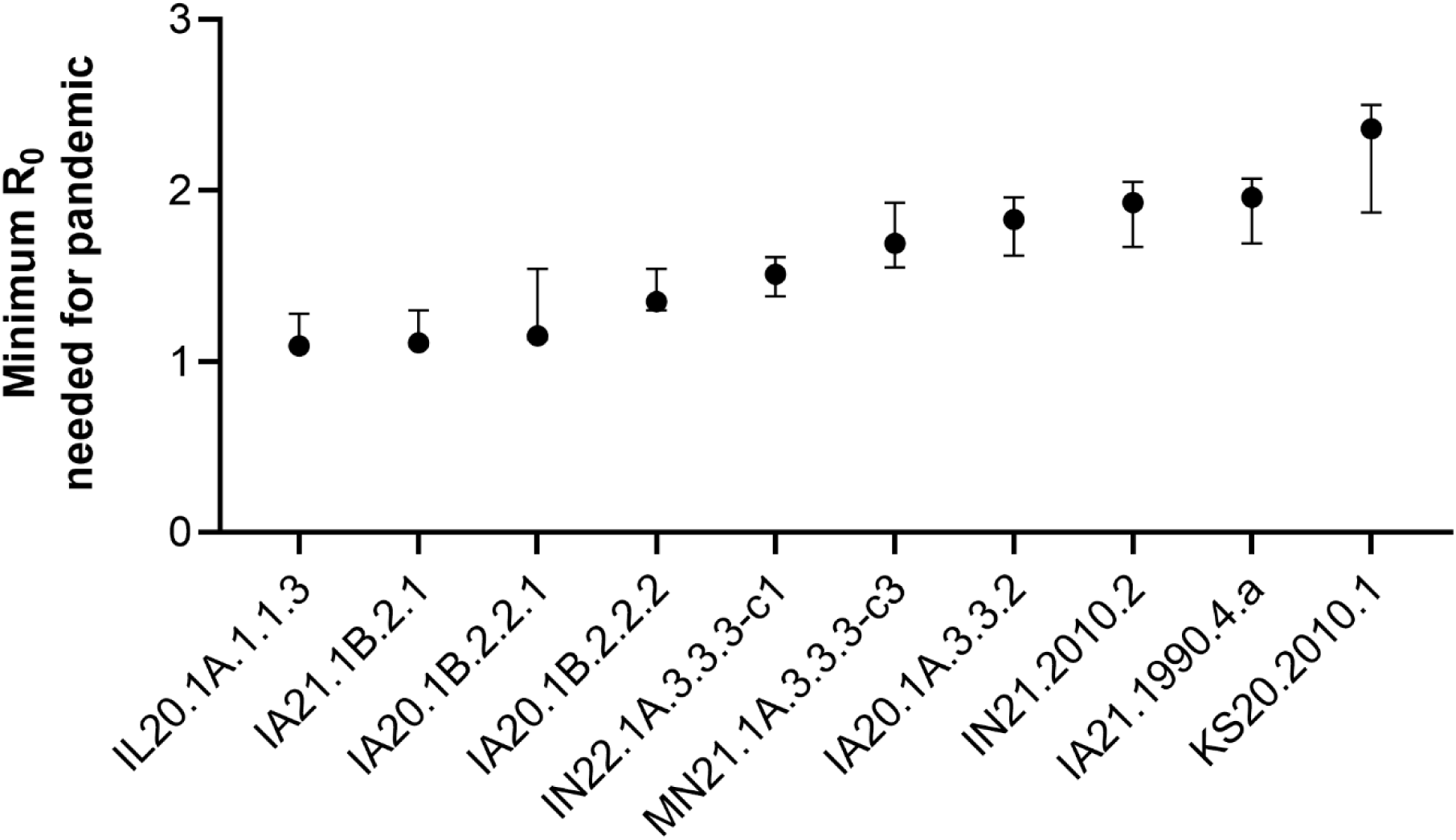
Minimum reproduction number (R₀) estimates required for the swine strains to cause a human pandemic derived from serological data. Error bars represent 95% confidence intervals. Strains are ordered by increasing minimum R₀ thresholds.

## Discussion

This study measured risk factors based on self-reported demographics of 4 human cohorts and applied two risk elements from the CDC’s Influenza Risk Assessment Tool to assess zoonotic risk of IAV in swine, population immunity and antigenic relatedness (*35*). Neutralizing antibodies were measured by HI assays against a panel of representative US swine IAV and human seasonal vaccine strains. Regardless of exposure to swine, all cohorts had gaps in population immunity against several strains of IAV circulating in U.S. swine, signaling vulnerability for both occupational groups and the general population to zoonotic transmission. In contrast to the hypothesis that occupationally exposed individuals are more likely to be infected with swine IAV and seropositive to swine strains, our findings revealed significantly lower odds of seropositivity to H1 1B and H3 swine strains for these individuals compared to the general population.

Demographic variables and vaccination status significantly influenced seropositivity to swine IAV. Occupational exposure to swine was associated with reduced seropositivity compared to general population cohorts for H1 1B (OR=0.23, p<0.001) and H3 lineages (OR=0.17, p<0.001) , likely reflecting lower seasonal influenza vaccination rates among occupationally exposed groups. This relationship was evident in both the reduced GMT and lower seroprevalence rates observed for human seasonal vaccine strains and contemporary swine strains among farm employees. The Philadelphia cohort displayed the highest overall antibody responses, likely reflecting the 100% uptake of the human seasonal influenza vaccine , and measured after vaccination at the peak of antibody response. Additional demographic factors with significant associations included birth cohort, with individuals born after 1977 showing higher seropositivity (H1 1A OR=1.94, p=0.038, H1 1B OR=3.47, p=0.002, and H3 OR=2.35, p=0.036), and female sex, which was associated with increased H3 seropositivity (OR=3.49, p=0.004). Swine farm employees displayed low vaccination rates at only 33.3%, 18 percentage points less than the national coverage estimates for the 2021–22 season (51.4%) (*36*).

Vaccination was key in inducing correlates of protection, H1 1A (OR=5.30, p<0.001), H1 1B (OR=5.48, p<0.001), and H3 (OR=14.44, p<0.001), where there was a significant increase in seropositivity for vaccinated compared to unvaccinated groups. Strain-specific analyses revealed vulnerabilities, with occupational groups showing significantly lower seropositivity to H1 1B strains IA21.1B.2.1 and IA20.1B.2.2.1 (OR=0.22-0.24, p<0.001) and H3 strain KS20.2010.1 (OR=0.06, p<0.001). Individuals lacked protective immunity against IL20.1A.1.1.3, IN22.1A.3.3.3-c-1, IA21.1B.2.1, and IA20.1B.2.2.2, indicating increased zoonotic risk for these endemic swine strains.

Gaps in seropositivity were further supported by the magnitude the swine strains differed antigenically from human vaccine strains. While antibody responses of many individuals from occupational and general population cohorts were within an 8-fold range of representative human seasonal vaccine strains, one swine strain showed significant antigenic distance, suggesting a potential mismatch between circulating swine viruses and existing human immunity. The representative clade 1A.1.1.3 had the highest significant antigenic distance from the H1 human seasonal vaccine strain (3.62AU), signaling a need for pandemic preparedness vaccine as the human seasonal vaccine likely provides little cross-protection from this swine H1 clade.

Four strains representing those circulating in the U.S. swine population demonstrated minimum R₀ thresholds below 1.46, the R₀ of the most recent 2009 IAV pandemic which notably originated from swine. Two swine strains, IL20.1A.1.1.3 (R₀ threshold= 1.09) and IA21.1B.2.1 (R₀ threshold = 1.11), were estimated to achieve pandemic spread with lower inherent transmissibility than any recorded influenza pandemic (*34*). These two H1 clades were detected consistently prior to 2021 (*19*) and represented 26.3% of the swine IAV detections across multiple US states during the study period. If these swine strains acquired moderate transmissibility in humans, existing population immunity would not be sufficient and pandemic spread could be established.

Although the individuals in this study were sampled from specific participating groups, and not a random population sample, patterns of vulnerability were identified. Individuals occupationally exposed to swine, such as Veterinarians and Farm Employees, were significantly less likely to have protective antibody levels against contemporary IAV in swine. This group exhibited lower vaccination rates and reduced seropositivity, particularly swine farm employees, underscoring a heightened risk of zoonotic infection. These findings demonstrate that immunity profiling can identify high-risk populations and potential pandemic threats before widespread transmission occurs. Risk analysis must be paired with preventative measures to reduce the risk of IAV infections in people and protect public health. At-risk populations working with swine should be targeted for promoting human seasonal IAV vaccine uptake to reduce zoonotic risk, with the added benefit of reducing transmission of human seasonal IAV to swine (*37*). Swine strains representing HA clades 1A.1.1.3, 1A.3.3.3-c-1, 1B.2.1, and 1B.2.2.2 demonstrated elements of increased pandemic risk, including low population immunity, lack of cross-reactivity to human seasonal vaccine strains, and low thresholds of required human transmissibility. To better protect swine-exposed individuals and the general population, candidate vaccine viruses should prioritize the inclusion of these strains during sequence selection for future pandemic preparedness vaccine development.

## Data Availability

All data produced in the present work are contained in the manuscript.

https://github.com/flu-crew/datasets

## Acknowledgements

We acknowledge Katharine Young for her contributions to the laboratory testing and Blake Inderski for management of the USDA IAV in swine surveillance database used for strain selection. We thank Sara Vogelaar for blood collection from veterinarians and farm employees, and Ben Cowling and Nancy Leung for their contribution of the sera samples and meta-data from the EPI-HK study, as well as comments during the preparation of this manuscript. The “<u>E</u>valuating <u>P</u>opulation Immunity in <u>H</u>ong <u>K</u>ong” (EPI-HK) study was funded in part by the Theme-based Research Scheme under project no. T11-712/19-N from the Research Grants Council from the University Grants Committee.

## Biographical Sketch

Celeste Snyder is a PhD student with the Virus and Prion Unit at the USDA National Animal Disease Center and Iowa State University. She studies zoonotic diseases at the human–animal interface, with research interests in public health and the application of a One Health approach.

## Address for Correspondence

Amy L. Baker, DVM, PhD, Research Veterinary Medical Officer, Lead Scientist Virus and Prion Research Unit, National Animal Disease Center USDA-Agricultural Research Service 1920 Dayton Avenue, Ames, IA. 50010, Phone: 515-337-7557

