## Appendix for "Occupationally exposed and general population antibody profiles to influenza A viruses circulating in swine as an indication of zoonotic risk"

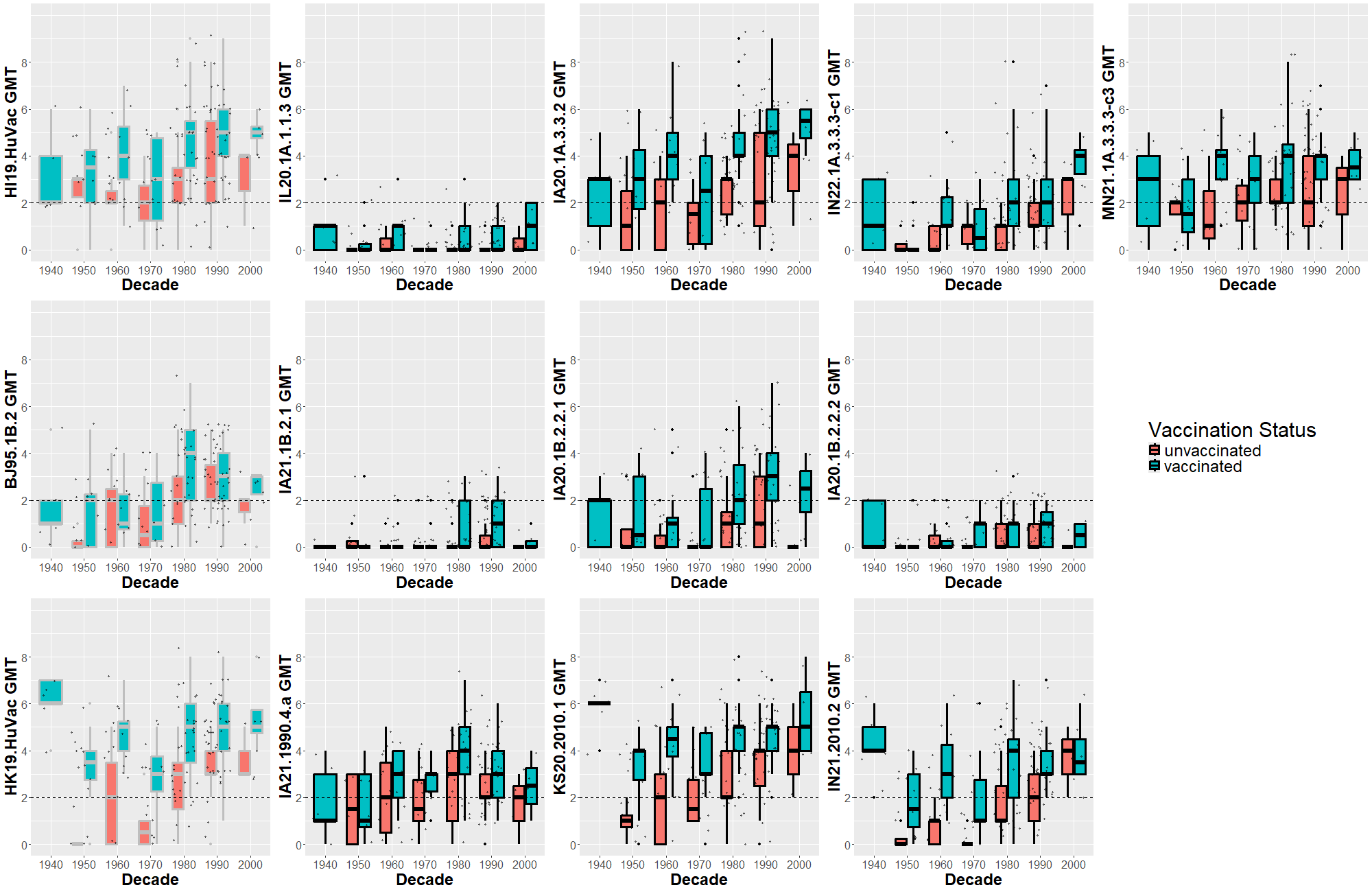

Appendix Figure 1. Antibody titers of all tested strains compared by vaccination status and decade of birth. Box and whisker plot of the geometric mean fold change in HI log_2_ titer against the swine strain compared to each respective participant’s HuVac HI titer within each cohort. Antigens are organized by H1 1A, H1 1B, and H3. Orange box plots represent unvaccinated individuals, and blue represents vaccinated individuals. Human seasonal vaccine strain boxplots are outlined in grey in the lefthand column.

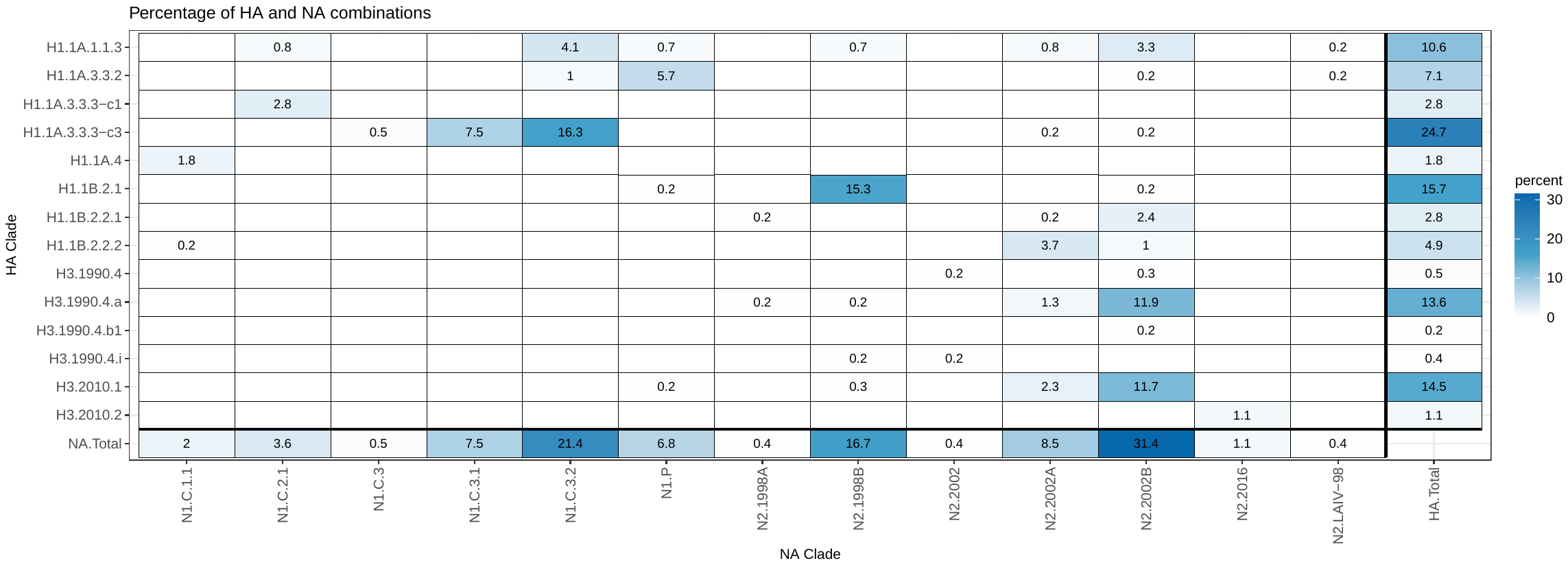

Appendix Figure 2. Heat map of percentage of hemagglutinin and neuraminidase combinations between July 2021 and June 2022.

Appendix Table 1. Odds ratios and 95% confidence intervals for associations with seropositivity to influenza A virus H1 and H3 lineages in swine. Statistically significant p-values (<0.05) are highlighted in bold.

| Variable | H1 1A | | H1 1B | | H3 | |
| --- | --- | --- | --- | --- | --- | --- |
|  | OR (95% CI) | p-value | OR (95% CI) | p-value | OR (95% CI) | p-value |
| General population cohorts | 1.00 (Referent) | - | 1.00 (Referent) | - | 1.00 (Referent) | - |
| Occupational exposure cohorts | 0.96 (0.53-1.73) | 0.899 | 0.23 (0.11-0.48) | **<0.001** | 0.17 (0.05-0.44) | **<0.001** |
| Born 1946-1977 | 1.00 (Referent) | - | 1.00 (Referent) | - | 1.00 (Referent) | - |
| Born 1978-2003 | 1.94 (1.04-3.66) | **0.038** | 3.47 (1.52-9.06) | **0.002** | 2.35 (1.06-5.22) | **0.036** |
| male | 1.00 (Referent) | - | 1.00 (Referent) | - | 1.00 (Referent) | - |
| female | 1.65 (0.91-3.01) | 0.098 | 1.92 (0.99-3.79) | 0.054 | 3.49 (1.48-9.32) | **0.004** |
| Non-vaccinated | 1.00 (Referent) | - | 1.00 (Referent) | - | 1.00 (Referent) | - |
| Vaccinated | 5.30 (2.56-11.32) | **<0.001** | 5.48 (2.15-17.17) | **<0.001** | 14.44 (5.33-47.30) | **<0.001** |

Appendix Table 2. Odds Ratios and 95% Confidence Intervals for Seropositivity to Individual H1 1A Swine Influenza A Virus Strains. Statistically significant p-values (<0.05) are highlighted in bold.

| Variable | IL20.1A.1.1.3 | | IA20.1A.3.3.2 | | IN22.1A.3.3.3-c1 | | MN21.1A.3.3.3-c3 | |
| --- | --- | --- | --- | --- | --- | --- | --- | --- |
|  | OR (95% CI) | p-value | OR (95% CI) | p-value | OR (95% CI) | p-value | OR (95% CI) | p-value |
| General population cohorts | 1.00 (Referent) | - | 1.00 (Referent) | - | 1.00 (Referent) | - | 1.00 (Referent) | - |
| Occupational exposure cohorts | 0.54  (0.10-2.34) | 0.409 | 0.87  (0.44-1.69) | 0.681 | 0.84 (0.46-1.54) | 0.581 | 1.36  (0.72-2.59) | 0.348 |
| Born  1946-1977 | 1.00 (Referent) | - | 1.00 (Referent) | - | 1.00 (Referent) | - | 1.00 (Referent) | - |
| Born  1978-2003 | 0.75  (0.17-4.01) | 0.715 | 1.48  (0.73-2.96) | 0.271 | 4.05 (1.94-9.18) | **<0.001** | 2.31  (1.18-4.51) | **0.015** |
| male | 1.00 (Referent) | - | 1.00 (Referent) | - | 1.00 (Referent) | - | 1.00 (Referent) | - |
| female | 3.48  (0.75-26.88) | 0.115 | 1.73  (0.88-3.49) | 0.113 | 1.53 (0.84-2.80) | 0.169 | 1.17  (0.62-2.25) | 0.630 |
| Non-vaccinated | 1.00 (Referent) | - | 1.00 (Referent) | - | 1.00 (Referent) | - | 1.00 (Referent) | - |
| Vaccinated | 4.32  (0.76-109.90) | 0.108 | 4.09  (1.82-9.50) | **0.001** | 3.31 (1.58-7.34) | **0.001** | 2.71  (1.22-6.09) | **0.014** |

Appendix Table 3. Odds Ratios and 95% Confidence Intervals for Seropositivity to Individual H1 1B Swine Influenza A Virus Strains. Statistically significant p-values (<0.05) are highlighted in bold.

| Variable | IA21.1B.2.1 | | IA20.1B.2.2.1 | | IA20.1B.2.2.2 | |
| --- | --- | --- | --- | --- | --- | --- |
|  | OR (95% CI) | p-value | OR (95% CI) | p-value | OR (95% CI) | p-value |
| General population cohorts | 1.00 (Referent) | - | 1.00 (Referent) | - | 1.00 (Referent) | - |
| Occupational exposure cohorts | 0.22 (0.08-0.53) | **<0.001** | 0.24 (0.13-0.44) | **<0.001** | 1.05 (0.45-2.48) | 0.903 |
| Born 1946-1977 | 1.00 (Referent) | - | 1.00 (Referent) | - | 1.00 (Referent) | - |
| Born 1978-2003 | 4.80 (1.59-21.60) | **0.004** | 2.69 (1.41-5.28) | **0.003** | 1.04 (0.43-2.72) | 0.928 |
| male | 1.00 (Referent) | - | 1.00 (Referent) | - | 1.00 (Referent) | - |
| female | 1.64 (0.74-3.69) | 0.220 | 1.97 (1.10-3.56) | **0.023** | 0.70 (0.29-1.62) | 0.403 |
| Non-vaccinated | 1.00 (Referent) | - | 1.00 (Referent) | - | 1.00 (Referent) | - |
| Vaccinated | 5.17 (1.68-23.48) | **0.003** | 3.42 (1.67-7.34) | **0.001** | 2.30 (0.86-7.42) | 0.100 |

Appendix Table 4. Odds Ratios and 95% Confidence Intervals for Seropositivity to Individual H3 Swine Influenza A Virus Strains. Statistically significant p-values (<0.05) are highlighted in bold.

| Variable | IA21.1990.4.a | | KS20.2010.1 | | IN21.2010.2 | |
| --- | --- | --- | --- | --- | --- | --- |
|  | OR (95% CI) | p-value | OR (95% CI) | p-value | OR (95% CI) | p-value |
| General population cohorts | 1.00 (Referent) | - | 1.00 (Referent) | - | 1.00 (Referent) | - |
| Occupational exposure cohorts | 1.16 (0.61-2.22) | 0.651 | 0.06 (0.00-0.29) | **<0.001** | 0.14 (0.06-0.31) | **<0.001** |
| Born 1946-1977 | 1.00 (Referent) | - | 1.00 (Referent) | - | 1.00 (Referent) | - |
| Born 1978-2003 | 3.04 (1.55-6.00) | **0.001** | 2.10 (0.78-5.59) | 0.140 | 2.31 (1.18-4.51) | **0.015** |
| male | 1.00 (Referent) | - | 1.00 (Referent) | - | 1.00 (Referent) | - |
| female | 1.39 (0.72-2.69) | 0.325 | 5.03 (1.58-23.22) | **0.005** | 4.22 (2.08-9.11) | **<0.001** |
| Non-vaccinated | 1.00 (Referent) | - | 1.00 (Referent) | - | 1.00 (Referent) | - |
| Vaccinated | 2.36 (1.08-5.19) | **0.032** | 13.22 (4.02-62.48) | **<0.001** | 5.85 (2.79-12.70) | **<0.001** |

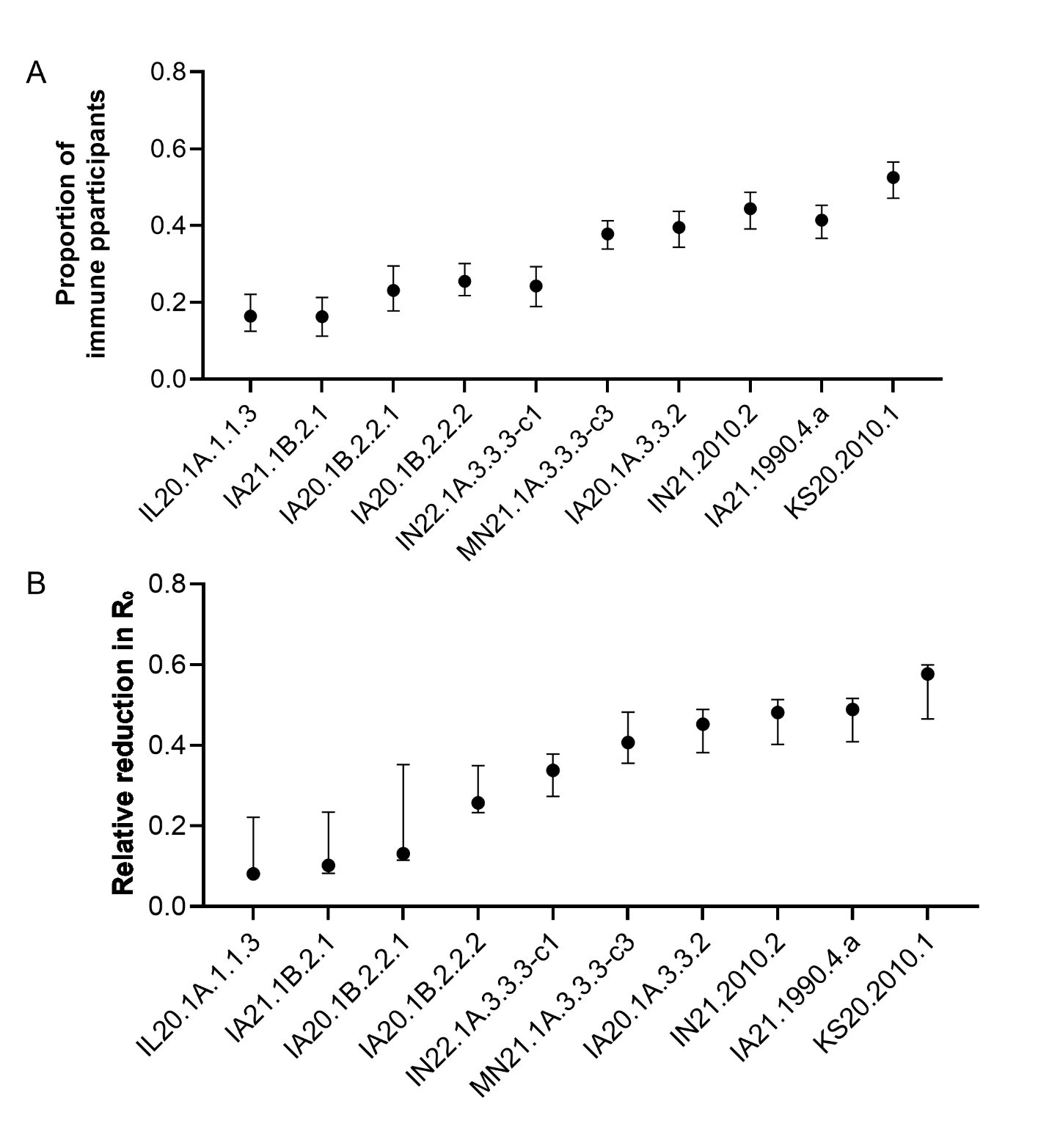

Appendix Figure 3. Reproduction number modeling with serological data for all participating individuals. (A) Proportion of immune participants (B) Relative reduction in R_0_. Error bars represent 95% confidence intervals. Strains are ordered by increasing minimum R₀ thresholds.
